# Machine Learning in Psychiatric Health Records: A Gold Standard Approach to Trauma Annotation

**DOI:** 10.1101/2025.03.09.25323272

**Authors:** Bruce Atwood, Eben Holderness, Marc Verhagen, Ann K Shinn, Philip Cawkwell, Hudson Cerruti, James Pustejovsky, Mei-Hua Hall

## Abstract

Psychiatric electronic health records present unique challenges for machine learning due to their unstructured, complex, and variable nature. This study aimed to create a gold standard dataset by identifying a cohort of patients with psychotic disorders and posttraumatic stress disorder, (PTSD), developing clinically-informed guidelines for annotating traumatic events in their health records and to create a gold standard publicly available dataset, and demonstrating the dataset’s suitability for training machine learning models to detect indicators of symptoms, substance use, and trauma in new records. We compiled a representative corpus of 200 narrative heavy health records (470,489 tokens) from a centralized database and developed a detailed annotation scheme with a team of clinical experts and computational linguistics. Clinicians annotated the corpus for trauma-related events and relevant clinical information with high inter-annotator agreement (0.715 for entity/span tags and 0.874 for attributes). Additionally, machine learning models were developed to demonstrate practical viability of the gold standard corpus for machine learning applications, achieving a micro F1 score of 0.76 and 0.82 for spans and attributes respectively, indicative of their predictive reliability. This study established the first gold-standard dataset for the complex task of labelling traumatic features in psychiatric health records. High inter-annotator agreement and model performance illustrate its utility in advancing the application of machine learning in psychiatric healthcare in order to better understand disease heterogeneity and treatment implications.

## INTRODUCTION

Psychotic and mood disorders are among the most disabling conditions worldwide[1–5], yet there is considerable clinical heterogeneity among patients. As a result, effective personalized treatments require a better understanding of this broad diversity in patient clinical histories. Among the different factors contributing to this heterogeneity, childhood trauma stands out as an under-recognized source and a major risk for the development of severe psychiatric disorders.[6–11] Among individuals with severe psychiatric disorders, the prevalence of childhood sexual, physical, and emotional abuse is significant at 26%, 39% and 34%, respectively,[12] and the average rate of all forms of trauma in adult females is 69% and in males 59%.[10] Substantial evidence indicates that individuals exposed to childhood physical or sexual abuse are 2-3 times more likely to develop a psychiatric disorder later in life, including psychosis.[6, 13–16] Research shows that childhood traumas impact critical windows of brain development and can trigger the onset of psychosis and other psychiatric disorders. In addition, among patients with psychotic and mood disorders, childhood trauma influences psychopathology, leading to more severe symptoms, higher rate of relapses or rehospitalization, higher rates of substance abuse, higher rates of treatment resistance, and poorer functioning.[11–13, 17–20] Although evidence clearly indicates that childhood trauma contributes to psychiatric risk and poor treatment outcome, large-scale computational approaches to stratify subpopulations, extract trauma features (e.g., frequency, type), and examine the links or the impact of trauma features on psychopathology and treatment outcome have yet to be developed.

The solutions to this major gap require leveraging computational methods over large data, human expertise, and the transdiagnostic approach (Research Domain Criteria [RDoC] framework operating on dimensional conceptualizations of mental illnesses) to provide clinical insights and utilize large data for accurate patient stratification. Such explainable machine learning (ML)[21] knowledge is the key to guide development of new therapeutic targets or interventions. Electronic health records (EHRs) contain structured and unstructured data over a patient’s treatment history. Unstructured data are richly detailed and add valuable contextual information but require additional processing to extract data elements suitable for aggregate analyses. Manual chart review[22, 23] is an impractical approach and is not cost-effective. Natural language processing (NLP) tools using EHRs have been developed,[24,25] which mine EHR data in a manner that is not only cost efficient, scalable and systematic, but could improve clinical precision by accurately classifying or stratifying patient subpopulations. For these tools to work effectively over EHR data, however, there needs to be an initial recognition of what textual aspects of the EHR help in classifying these populations. This is accomplished through annotation.

The goal of the annotation is to mark up (tag) clinical information present in the EHRs (e.g., events, symptoms, temporal expressions, etc.) in order to train NLP models to automatically extract this information from clinical text. The Informatics for Integrating Biology and the Bedside (i2b2) project is one example of a major effort which has successfully developed annotation guidelines, gold standard datasets, and resources for use by the medical community.[26–28] However, i2b2 focuses on medical related conditions (e.g., diabetes) and concepts. To our knowledge, there is currently no “computational version of chart reviews” tool that systematically extracts trauma related features and stratifies patients into subgroups (with or without childhood trauma history). In addition, annotation guidelines representing psychiatric patients and trauma-related features have yet to be developed. A gold standard corpus (i.e., annotated EHR text dataset) specifically tailored to identify trauma-related events, risk factors, symptoms, and relations between them does not exist. The lack of such resources is a major barrier in the progress for the development of accurate ML classifiers and prediction models in psychiatry.

The contributions of this study are to: 1) develop an annotation scheme and clinically informed guidelines for labeling trauma related linguistic features; 2) create training and testing corpora of psychiatric patient EHRs and establish a publicly available gold standard dataset, “Trauma Enriched Psychiatric Corpus” (TEPC); and 3) use the TEPC to train and evaluate ML models. To our knowledge, the set of resources developed in this study is the first to detect/identify linguistic features of trauma related concepts in patients with psychosis and mood disorders.

## MATERIALS AND METHODS

### Dataset

EHRs were sourced from the Research Patient Data Registry (RPDR),[29] a centralized regional data repository which contains discharge summaries, encounter notes, and visit notes for over 7 million patients across all institutions in the Mass General Brigham healthcare network from 1999 onwards. Using the RPDR query tool, we aggregated all EHRs of patients who met three criteria:

1. Diagnoses of both PTSD and a psychotic disorder (schizophrenia, bipolar, schizotypal, delusional, or other non-mood psychotic disorder) at some point in patient history.
2. Between 18 and 65 years of age at time of query (Oct. 2021).
3. At least one note related to psychotherapy, symptoms referent to mental disorders, or social counseling.

This search yielded approximately 55000 notes over 3515 patients. The dataset was designed to ensure broad applicability across different ages and genders, focusing on a rich density of annotatable content. To this end, we first quantified the presence of traumatic event-related keywords (as detailed below) in each clinical note from the RPDR query. The notes were then categorized by age and gender, selecting the ones with the highest frequency of these keywords in equal proportions from each demographic group. 200 narrative heavy clinical notes were selected. These notes contained significantly more annotatable content than typical EHRs, with an average of 2353 tokens per document compared to 617 and 709 tokens per document in the 2014 i2b2/UTHealth [28] and MIMIC-III [30] datasets, respectively. The sex breakdown was 103 females to 97 males, with a mean age of 35.8 and a median age of 35.

### De-identification and pre-annotation processing

The 200 clinical notes underwent a de-identification process to remove all personally identifiable information in accordance with HIPAA guidelines. Each document was manually inspected to eliminate any of the 18 identifiers specified under the HIPAA Privacy Rule’s Safe Harbor method [38].To maintain the distinction between different individuals mentioned in the notes, names were replaced with unique labels, such as converting “John Doe” to “<patient>” and “Dr. Alice Bob” to “Dr. <person1>”.

Before manual annotation, the documents were pre-annotated automatically for explicit references to trauma-related symptoms, events, and substance abuse. This pre-annotation schema, adapted from Jackson et al.[31], is detailed in the Pre-annotation section of the additional materials. The purpose of this automatic pre-annotation was twofold: to lessen the workload of the annotators and to achieve high recall in the annotation process by systematically identifying all straightforward instances of the targeted categories (e.g., symptoms such as ‘self-injurious behavior’). Following this, the documents underwent manual annotation to identify and label more complex contents.

### Annotation scheme and guidelines

Annotation in this study involves labelling the clinical narratives with three primary types: entities, attributes, and relations. Entities are tied to specific text fragments and classified into various categories (for example, “emotional abuse” is labeled as an EVENT while “anxious” is labeled as a SYMPTOM). In the trauma annotation guidelines (TraumaML), entities would specifically focus on: traumatic events, perpetrators, symptoms, substances, and temporal frames that ground traumatic events within a context of time. Attributes provide additional details about an entity class (for instance, a symptom can be negated, as in “does not appear anxious,” to show the absence of that symptom). Relations link one entity to another, such as connecting an EVENT entity with a PERPETRATOR entity. (see annotation guideline for details)

The process of developing these annotation guidelines was based on the Model-Annotate-Model-Annotate (MAMA) cycle [32], as illustrated in Figure 1. In the initial stages of the project, draft guidelines were formulated using a preliminary set of categories for entities, relations, and attributes, chosen for their clinical relevance. These guidelines were then applied in a pilot annotation round. Subsequent discussions and revisions of the guidelines were informed by the outcomes of this pilot annotation.

**Figure 1.**
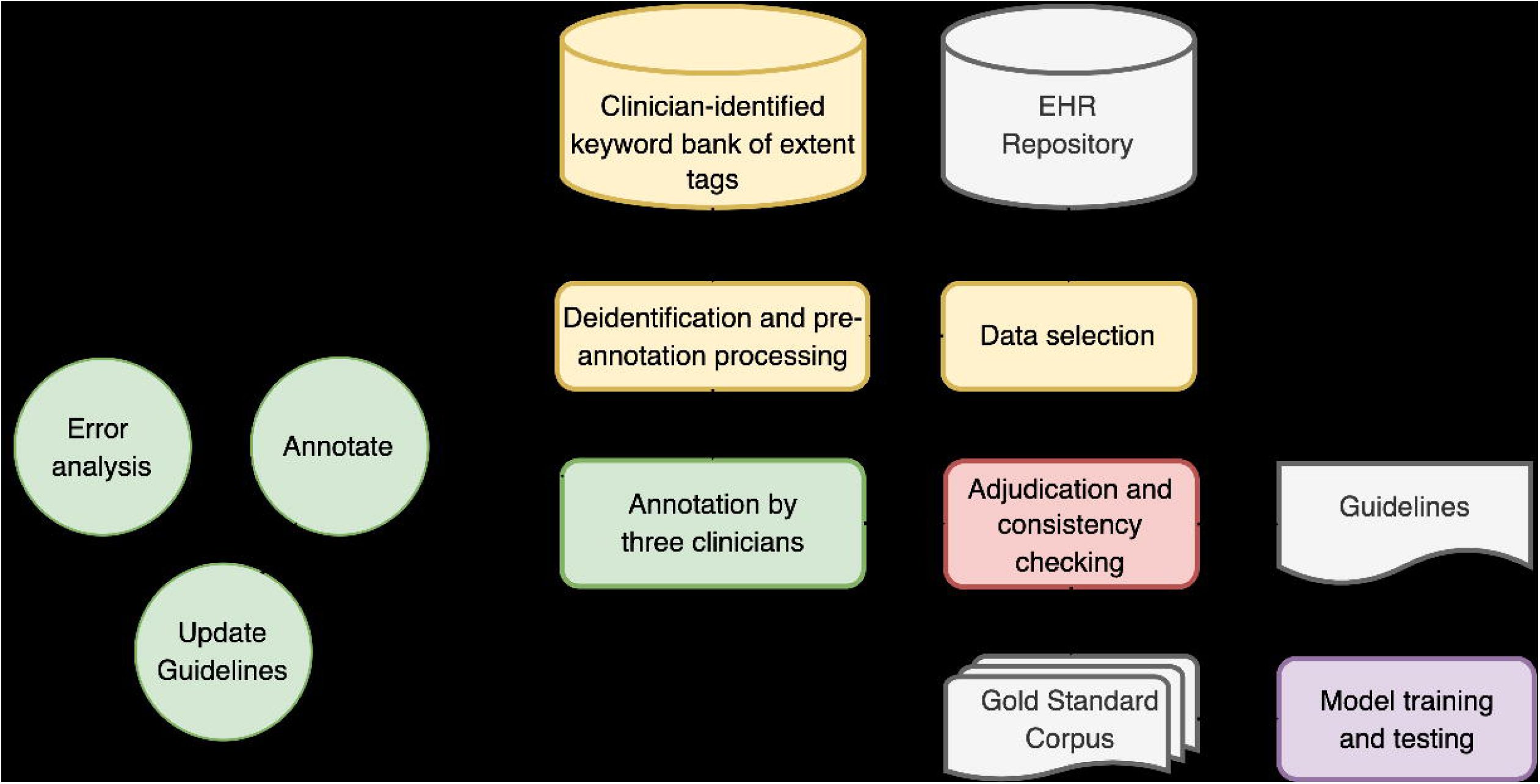
Graphical description of the project workflow. The yellow elements denote the dataset creation and preprocessing steps; green elements denote the annotation process; red denotes the revision stage; gray elements denote the inputs and outputs including the resulting Gold Standard trauma enriched psychiatric corpus; purple refers to the baseline models trained using the corpus.

Annotations were completed by a team of three domain experts (MHH, AS, PC) using the BRAT rapid annotation interface,[33] an open-source natural language annotation tool. Annotators first manually validated the automatically generated annotations. Relation tags were then created between pertinent span tags and assigned attribute labels to both the span tags and relations. An example is given in Figure 2.

**Figure 2.**
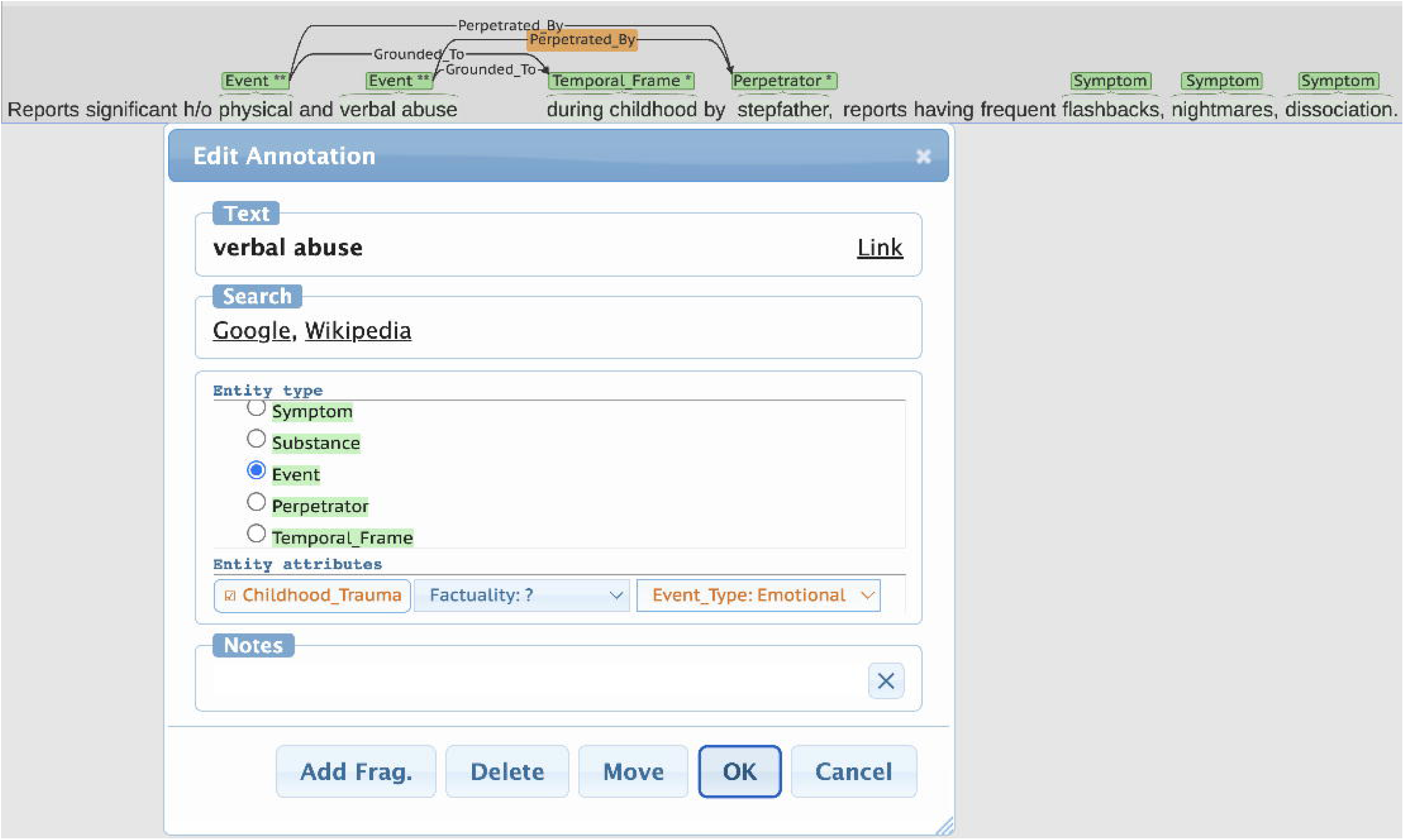
A screenshot of the BRAT annotation interface. When highlighting a span of text or drawing a relation between two annotated spans, a popup window is shown to the annotators where they may select the appropriate tag.

The Trauma annotation guidelines were iteratively refined by the domain experts in consultation with the team of NLP researchers, using a consensus-based approach, broadly following a set of guiding principles:

1. Annotators should approach the text with a neutral stance. They should not make inferences regarding psychiatric diagnoses, nor make assumptions that are not plainly made by the text itself. For example, “denies depressive mood, but appears tearful and sad.”
2. Annotations should maximize the pertinent clinical information. For example, “delusions with hyper-religious themes” instead of “delusions”.
3. Annotations should optimize model performance by maximizing replicability (reducing inter-rater disagreement) and minimizing the complexity of the machine learning task. For example, the span “suicide attempt by jump from a bridge”, “suicide attempt” is annotated.

Though the second and third principles are generally in contention, where an increase in clinical complexity leads to a more difficult modeling task, maximizing replicability is invariably beneficial. To this end, extensive error analyses were utilized throughout the process.

### Error analysis

The primary concern in developing an annotation guideline is consistency; the performance of any model depends foremost on a congruent set of patterns from which it may draw inferences. To this end, annotation discrepancies were analyzed and corrected at two levels: the intra-document scope through dual annotation (local error analysis) and at the inter-document scope through tag context (global error analysis).

### Local error analysis: mini batch dual annotation

At several points during the MAMA cycle, small batches of notes (26 notes in total) were annotated by two independent annotators rather than one. These 26 dually annotated notes allowed for a direct comparison between annotators, who then discussed disagreements with the goal of harmonizing annotation and fine-tuning the annotation guidelines. Inter-Annotator Agreement (IAA) scores were calculated over these 26 dually annotated documents using the pairwise F1 metric (defined as the harmonic mean of precision and recall), which were then averaged across each annotator pair to get a single score for each entity and relation. F1 score was chosen as it has a specificity of measurement that accounts for distinctions between false positives and false negatives.

For relation tags, two types of F1 scores were computed: a standard F1 and a ‘relaxed’ F1. The relaxed F1 score considered only those relation tags where the span tag constituents (the individual elements forming the relation) were in exact agreement between the annotators. This approach was applied to a total of 255 relation annotations. In contrast, the standard F1 metric was applied more broadly, encompassing all 403 relation annotations, including those where the span tag constituents did not match and were therefore considered to be in disagreement by default. This distinction between standard and relaxed F1 scores allowed for a more nuanced understanding of annotator agreement, particularly in the context of relation tagging.

Additionally, F1 scores for IAA as well as the model were calculated such that events and attributes were independent of each other, such that if a span was correctly deemed an event, but was misappropriated the emotional tag, this mistake was not penalized when calculating the error of the event entity tag, as it would be if the class event_emotional was treated as its own class.

### Global error analysis: left context of spans

Error analysis performed during dual annotation will find only local inconsistencies, i.e., differences in how annotators approach a specific text span, not whether the approach is consistent across documents. Therefore, it is also necessary to check for inconsistency at the global scope. This process focused particularly on the variation of tag extents, as extent is not as dependent on context as other features such as attributes.

To identify global inconsistencies, we developed a tool to analyze the left context of all annotated spans. First, we calculated the pointwise mutual information (PMI) between each tagged span and the token to its immediate left using the standard definition of PMI:

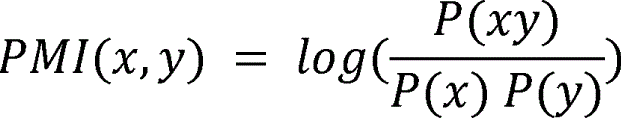

Where x is the token to the left, y the span, P(xy) the expected probability of the two occurring together, P(x) the probability of the token to left occurring and P(y) the probability of the span.

If the mutual information is high (>5) and the span and the token were annotated together somewhere in the corpus, we discussed which annotation extent was preferable and revised the guidelines accordingly. An example of the interface is presented in Figure 3.

**Figure 3.**
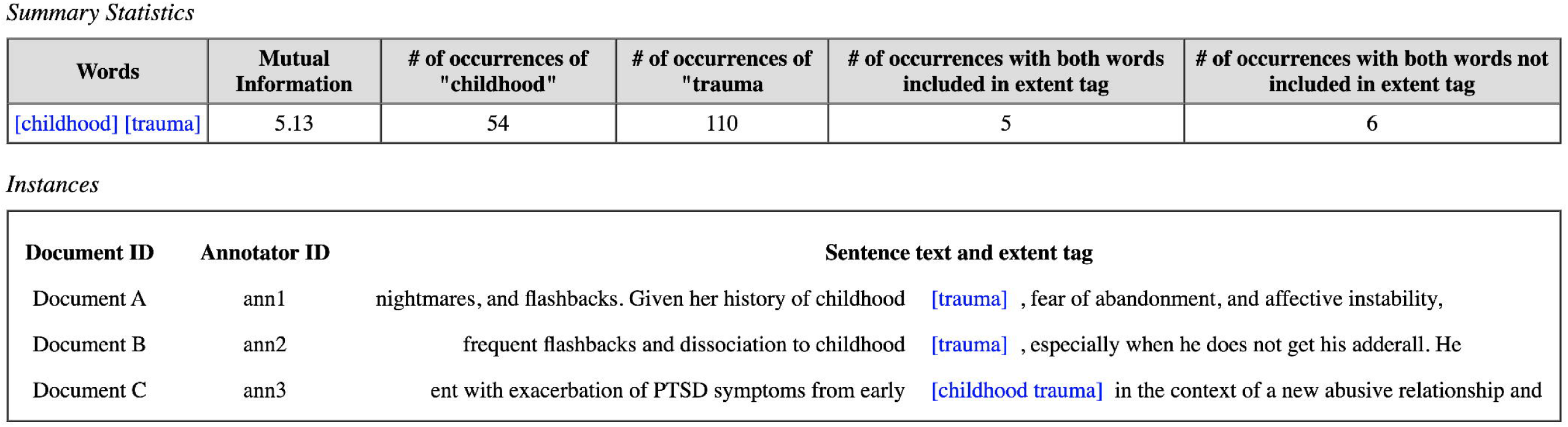
Analysis of annotations including the span “trauma” and preceding token “childhood.” A high mutual information score of 5.13 indicates potentially sufficient support of the combined annotation “childhood trauma” for use in annotation and modeling. In this example, across three documents A, B, and C one annotator annotates all of “childhood trauma”, while two other annotators only include “trauma.” These differences would not be found by comparing dually annotated notes as these notes were not dually annotated.

### Adjudication and Consistency Checking

When a consensus was reached regarding all conflicts and the annotation guidelines were finalized, the gold standard corpus (Trauma Enriched Psychiatric Corpus, TEPC) was considered complete. The dually annotated notes were adjudicated, resolving all discrepancies between individual annotators, and all other notes were updated to reflect the current version of the guidelines.

### Machine Learning Modeling

Baseline models were trained on the gold standard corpus to validate that the annotations are learnable and to serve as a foundation for future machine learning work. The modeling task is conceptually analogous to named entity recognition (NER), and is approached as a sequence-to-sequence token classification task, where the task of the model is to take in pre-tokenized input and label each token either as ‘O’ (signifying no label) or as one of the entity tags from the annotation specification.

The state of the art in token classification are transformer-based architectures, specifically BERT (Bi-directional Encoder Representations from Transformers) [34] and its variants. These models are pretrained on a large corpus and then may be finetuned on a smaller, labelled dataset relevant to the intended task. They create context-dependent embeddings of tokens, which may be then fed into a standard neural network for classification. This model structure is presented in Figure 4. In this study, the selected model was RoBERTa, a variant that has demonstrated superior performance over BERT in various tasks, notably in NER. Although RoBERTa retains the architectural framework of BERT, it distinguishes itself in several key aspects. Primarily, the training corpus size for RoBERTa is significantly larger, encompassing 160 GB of data, in contrast to the 16 GB corpus utilized for BERT. Additionally, RoBERTa and BERT differ in their approaches to pre-training loss functions and the settings of training hyperparameters [35].

**Figure 4.**
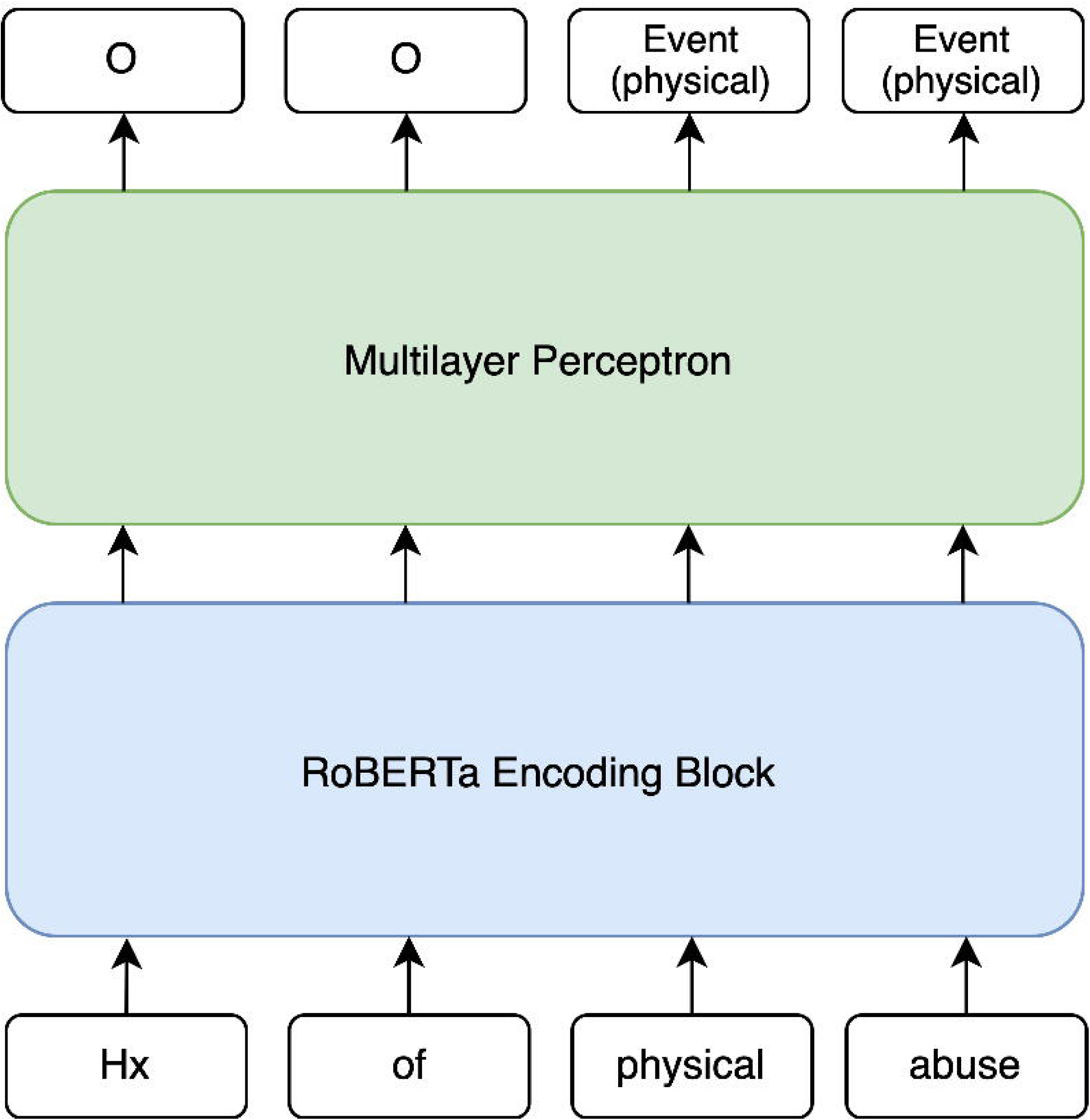
Representation of the model. The RoBERTa encoding block takes a sequence of word tokens as input and creates encodings for them, which are then piped into a standard multilayer perceptron for classification. Consecutive tokens labelled as entities of the same kind are considered to be a single entity (e.g., ‘physical abuse’).

In addition to the named entity recognition classifier that labelled spans, early versions of the baseline modelling paradigm featured a classifier that took each valid pair of tags from the span-level tagger and labelled the relationship between them, the details of this earlier version may be found in the supplementary materials section **Baseline model configuration and results** [see Additional File 1]. We found that the overall performance of the baseline model was not optimal and specific categories such as Perpetrator, Temporal_Frame performed poorly. In order to improve model performance, we made several fundamental changes to this earlier iteration with more training data. The relation classifier was discarded due to the low support of the entity tags (Temporal_frame and Perpetrator) grounding the relation tag, leading to modelling instability. Additionally, because RoBERTa has a maximum input constraint of 512 tokens, documents need to be segmented into chunks to be passed individually into the model. The length of these chunks in the baseline model was previously fixed to be a single parsed sentence. In the improved model configuration, we included the segmentation size as a hyperparameter in the hyperparameter sweep, under the hypothesis that constraining the context could negatively impact the classification performance.

The dataset was partitioned into training, development, and test sets, adhering to an 80:10:10 ratio. We employed a hyperparameter sweep to find values for learning rate, batch size and number of training epochs that optimized the F1 score of the model when evaluated on the development set. The best model was then evaluated on the held-out test set.

## RESULTS

### Annotation guidelines

TraumaML is designed for annotation of EHRs for patients with diagnosis of PTSD and psychotic and mood disorders. Detailed annotation guidelines are presented in the **Trauma Annotation guidelines** section of the supplementary materials [see Additional File 2].

The gold-standard guideline contains five entity tags and three relation tags. Below are the five entity tags that mark spans of text and the three relation tags that connect entity tags.

Entity Tags:

1. **Event**: The traumatic event that a patient has experienced.
2. **Perpetrator**: The perpetrator of the traumatic event.
3. **Symptom**: Symptom exhibited by the patient.
4. **Substance**: Any text span that gives information about a substance use disorder.
5. **Temporal_Frame**: A temporal grounding for the traumatic event. Relation Tags:
6. **Perpetrated_By**: Connects an event to a perpetrator.
7. **Grounded_To**: Connects an event to a temporal frame.
8. **Sub-Event**: Relation between an event and another event that it is part of.

For **Event** entity tags, we focused on three types of traumatic events that are interpersonal in nature (i.e., sexual, physical, and emotional abuse). Trauma events that were witnessed but not directly experienced by the patient (e.g., witnessing the death of a loved one) were categorized as “other.” Events that could be perceived as traumatic but that were not forms of interpersonal trauma (e.g., physical or psychological injury from a natural disaster or motor-vehicle accident) were marked as “other”. Trauma-related terms that reflect a medical condition (e.g., traumatic brain injury) were not annotated. (See the final guidelines for details.)

For **Symptom** entity tags, we annotated all symptoms, not just those that were documented by clinicians as being associated with specific trauma events. This is because trauma is associated with a wide range of psychopathology (psychosis, mood, anxiety), not only to posttraumatic stress symptoms (e.g., flashbacks, nightmares), and because symptoms may overlap across diagnostic conditions (e.g., sleep disturbance is seen across multiple psychopathologies). Additionally, we accounted for attributes like Negation, indicating the absence of a symptom, and Not Current, signifying symptoms occurring in the past.

**Substance** entity tags were annotated only if they reflected clinically defined substance abuse/dependence such as “alcohol use disorder,” or employed homologous language such as “complicated withdrawal,” “detox,” or “daily heroin use.” Text not directly indicative of abuse/dependence, such as positive results of toxicology screens or other unspecified drug/alcohol use, was not considered, and no differentiation between current and past substance abuse was made.

### Gold-standard “Trauma Enriched Psychiatric Corpus” (TEPC)

Table 1 shows the distribution of the gold standard TEPC corpus, as well as the inter-annotator agreement results from the 26 dually annotated documents. Overall, the corpus contains 15 013 entity span tags and 1136 relation tags. Most frequent span tags are **Symptom** (11,568), followed by **Event** (1357), then **Substance** (1147).

**Table 1.** Annotation Results.

|  | Count | Mean IAA F1 Score |
| --- | --- | --- |
| Documents | 200 | - |
| Tokens | 470 489 | - |
| **Entity Tags** | 15 013 | - |
| **Event** | 1357 | 0.68 |
| Childhood | 372 | 0.81 |
| Physical | 289 | 0.82 |
| Emotional | 99 | 0.80 |
| Sexual | 421 | 0.88 |
| **Symptom** | 11 568 | 0.73 |
| Negation | 2449 | 0.94 |
| Not_Current | 1191 | 0.82 |
| **Substance** | 1147 | 0.72 |
| **Temporal_Frame** | 517 | 0.51 |
| Time-Of-Life | 138 | 0.87 |
| Age | 86 | 0.82 |
| Date | 87 | 0.72 |
| Period | 203 | 0.59 |
| **Perpetrator** | 424 | 0.63 |
| Family Member | 207 | 1.00 |
| Other-Known | 110 | 0.81 |
| Other-Unknown | 51 | 0.59 |
| Partner | 56 | 0.87 |
| **Relation Tags** | 1136 | - |
| **Grounded_To** | 585 | 0.60 (0.94) |
| **Perpetrated_By** | 496 | 0.64 (0.95) |
| **Sub-Event** | 55 | 0.37 (0.50) |

Note: distribution of gold standard TEPC corpus and pairwise inter-annotator agreement (IAA) scores. IAA scores were calculated for each entity class. For relation tags, the relaxed fit F1 scores are provided in parentheses.

### Model performance

Per-label results for the entity classification model are reported in Table 2, using the optimal hyperparameters that were identified through the hyperparameter sweeping described in the Methods section. The model achieved a macro average F1 score of 0.76.

**Table 2.** Per-label results of entity classification model.

|  | support | precision | recall | f1-score |
| --- | --- | --- | --- | --- |
| **Event** | 146 | 0.75 | 0.73 | 0.74 |
| Childhood | 34 | 0.65 | 0.71 | 0.68 |
| Physical | 17 | 0.65 | 0.76 | 0.70 |
| Emotional | 10 | 0.88 | 0.70 | 0.78 |
| Sexual | 96 | 0.90 | 0.85 | 0.88 |
| **Symptom** | 1086 | 0.78 | 0.76 | 0.77 |
| Negation | 245 | 0.86 | 0.83 | 0.85 |
| Not Current | 77 | 0.77 | 0.70 | 0.73 |
| **Substance** | 110 | 0.64 | 0.75 | 0.69 |
| **Micro Avg** | 1821 | **0.78** | **0.77** | **0.78** |
| **Macro Avg** | 1821 | **0.76** | **0.75** | **0.76** |

## DISCUSSION

To the best of our knowledge, this study represents the first investigation of trauma-related features in a large and representative cohort of psychiatric electronic health records. In this study we developed clinically informed guidelines for annotating these patient health records for instances of traumatic events, created a gold standard publicly available dataset, and demonstrated that the data gathered using this annotation scheme is suitable for training a ML model to identify indicators of trauma in novel (unseen) health records. The preprocessing steps, guidelines, as well as the code for NLP models, are available as a repository on our GitHub group [37]. We identified five entity tags (**Event**, **Perpetrator**, **Symptom**, **Substance**, and **Temporal_Frame**) and three relational tags (**Perpetrated_By**, **Grounded_To**, and **Sub-Event**) as important for modeling trauma-related linguistic features in psychiatric EHR data.

Identifying symptoms and trauma-related features in the mental health domain is challenging because psychiatric clinical narratives have considerable variability, both because of the heterogeneity of patient presentations as well as heterogeneity in the way that different clinicians may document clinical events. Nuance is intrinsic to the annotation process of psychiatric EHRs, and in many cases there is no obvious justification for a correct way to annotate a span. The following discussion aims to shed light on the consistent points of disagreement and complexity in the annotation process and how we navigated these challenges. Our efforts serve as a guide for future annotation endeavors, providing insight into how to effectively identify symptoms and trauma-related features while accounting for the inherent nuance and variability of clinical narratives.

We structured our annotation process around three key principles: (1) take a neutral approach to the text; (2) capture as much relevant clinical information as possible; and (3) enhance model performance by reducing complexity. There is a trade-off between principles #2 and #3, as adding more details (e.g. additional types of annotations or annotations that tag more detailed information) makes the learning task harder and reduces the number of training examples for more specific descriptions. The guidelines were crafted based on the project’s scope and the model’s capabilities to draw meaningful insights from a fixed training corpus size. To demonstrate how these principles were applied in the guidelines, we discuss the resolution of several key disagreements that arose during the annotation process.

In some cases, annotators found it difficult to decide the truthfulness/credibility of a traumatic event, due to the acute state of psychosis of a patient (e.g., delusional). In these instances, events were assigned the default attribute, factual. Annotators marked attributes maybe or unlikely only when the sentential context is unambiguous that the event in question is uncertain or dubious. Thus, “sexual assault” in the span “possible sexual assault” was attributed as maybe, but in the line “pt noted that she had been assaulted…” further in the same document, the span “assaulted” was attributed as factual. This conservative approach is an exemplification of principle #1, where the text is taken at face value based on the current limitations of language models to infer nuanced context, especially over prolonged reading frames. Substance use was similarly annotated, where unspecified drug/alcohol use was ignored when the clinician did not specify it as a clinical problem.

Additionally, many questionable situations during annotation required balancing annotation complexity with modeling complexity. For instance, to establish consistency on span tag extents, the extent of a span tag was determined as the minimal extent to capture the information pertinent to the study. An example of this would be deciding to annotate “self-injury” as a symptom only, rather than the full span “scratch herself superficially as self-injury.” Although this decision results in the loss of clinical context, the inclusion of additional information would be deemed extraneous if the training data does not provide sufficient support for a model to draw an inference about the symptom in question. In this way principles #2 and #3 are balanced. Importantly, this balance is contingent on the scope of the study. Because we were focused specifically on childhood trauma, we decided to only annotate “trauma” within the context “military trauma”, as the gains in specificity do not offset the loss in generalizability.

To simplify the annotation task and still capture specific temporal links between temporal expressions such as dates, and available events ({sexual abuse} in {2002}), we created the **Temporal_Frame** tag, consisting of 5 possible temporal type attributes: age, duration, time-of-life, major event, and date. The relation between the temporal frame and the event is expressed with the **Grounded_to** relation. We do not include other temporal relations such as defined by the Temporal Relation (TLINK) link described in the NIMH-THYME (Temporal History of Your Medical Events) guidelines as it is beyond the project scope. Although a framework of temporal relations to the onset of psychosis symptoms has been reported,[36] this framework focuses on psychosis symptom onset identification and is based on a limited set of symptom keywords which are not suitable to capture more complex linguistic variants. Developing temporal NLP systems in mental health records remains a challenge, due to the inherent complexity of the task. We plan to develop a psychiatric temporal relation annotation scheme and build temporal information extraction systems for psychiatric notes using a graph neural network method in the future.

### Modeling

Our entity classification model achieved a macro-average F1 score of 0.76 on the held-out test set, thus demonstrating the practical viability of the gold standard for machine learning applications. This performance metric is high considering the complexity of the task, which requires a broad range of annotations over unstructured and highly variable text.

This performance was achieved in part by reducing the scope of the model; the relation classifier component was removed, as well as the span-level entities ––namely, perpetrator and temporal frame–– that are specifically associated with relation tags. This choice was informed by the fact that modeling relations separately from span-level entities carries the risk of errors from the span-level model being transferred and amplified within the relation-level model, leading to compounded inaccuracies across different models. Addressing this issue in future research may involve adopting an integrated model that jointly identifies both entities and their relations, moving away from the sequential, pipeline approach employed in this study.

### Limitations and related future considerations

There are a few limitations that should be highlighted. First, given the complexity of psychiatric clinical notes and expertise required of annotators, performing annotation is time and resource expensive comparing to other domains (e.g., annotate recipes). This is a major limiting factor for scaling up the amount of annotated data. Second, among the annotated corpus, 26 notes were annotated by two annotators whereas the remaining 174 were not. However, these 174 singly annotated notes were retroactively updated to reflect changes to the guidelines that occurred while resolving discrepancies in the dually annotated notes. Third, TraumaML is designed for annotation of EHRs for patients with diagnosis of PTSD and psychotic and mood disorders. It doesn’t cover all psychiatric diagnoses (e.g., addiction). However, the annotation schema and guideline can be incorporated, expanded, and modified to suit for different psychiatric conditions that extend beyond the scope of this project.

## CONCLUSION

In this study we developed an annotation scheme and associated guidelines for marking indicators of trauma in psychiatric patient EHRs, undertook an annotation task with three domain expert clinicians, which has in turn, resulted in our creating a gold standard corpus for these indicators, the TEPC. Furthermore, we verified that the schema is learnable by ML algorithms by building a entity classification model that will be improved in future work for use in analyzing large volumes of unlabeled clinical data to investigate the relationship between trauma (including types of events, perpetrators, and when the trauma occurred) and clinical outcomes.

## Supporting information

TraumaAnnotationGuidelines-medrxiv

## ACKNOWLEDGMENTS

We thank funding agent NIMH for supporting this work. This work is supported by NIMH fund R21MH125076 (Mei-Hua Hall & James Pustejovsky, PIs)

## AUTHOR CONTRIBUTIONS

B.A. contributed to the data collection, methodology, software development, analysis, baseline modeling, and manuscript writing.

E.H. contributed to the data collection, methodology, software development, design of annotation task, analysis, baseline modeling, and manuscript writing.

M.V. contributed to design of annotation task, guideline development, analysis, and manuscript writing.

A.K.S. contributed to annotation task, guideline design and clinical expertise.

P.C. contributed to annotation task, guideline design and clinical expertise.

J.P. contributed to the conceptualization of the study, and methodology of the project.

M-H.H. contributed to the conceptualization of the study, methodology, annotation task, guideline development, manuscript writing, and provided resources and guidance on the overall direction of the project.

H. C. contributed to annotation task.

## DATA AVAILABILITY STATEMENT

Deidentified data is available at the National Data Archive (grant reference R21MH125076-01). Code used for pre-annotation and baseline modeling can be found on our GitHub page < https://github.com/TraumaML/annotation >. Trauma Enriched Psychiatric Corpus is available upon request.

## FIGURE LEGENDS

**Additional File 1**

Word Document [TP_Supplementary File_final.docx]

Supplementary Material

Supplementary information regarding pre-annotation, guideline development discussions, and baseline model and configurations

**Additional File 2**

PDF [TraumaAnnotationGuidelines.pdf]

Annotation guidelines

Guidelines that annotators developed over the course of the annotation phase of the project.

