## supplementary materials for "Machine Learning in Psychiatric Health Records: A Gold Standard Approach to Trauma Annotation"

**Pre-annotation**

For each category, we generated lists of regular expressions where any pattern matching a particular expression would be tagged as an instance of that class. For example, under the symptoms list, an expression might be

sleep* or slept [0-2_words] erratic*

which would capture phrases such as “sleep is erratic”, “sleeps erratically”, and “slept a little erratically” and annotate them as symptoms. The lists of expressions were iteratively trialed on documents and updated to reduce error. The final lists are included in the supplementary files.

The use of results from this stage requires oversight during the manual annotation stage as it is merely a tool to help the annotators, and erroneous annotations may still exist, such as “sleep is not erratic” without a negation tag. Within the pre-annotation script, it is necessary to account for sentence boundaries, i.e., in the span “… helped her sleep. Her erratic behavior…”, the sub-span “sleep. Her erratic” should not be annotated. The script must also allow for annotations that may span multiple lines, such as “sleeping[\n]erratically”.

**Guideline development discussions**

Many of these discussions focused on what exact span should be annotated. The general rule of thumb is to annotate the extent that is most informative and clinically relevant. For example, the “shaking” is the most important part in “her arm began shaking” and adding that it was the arm that was shaking does not add a lot of information or clinical relevance. Two other general rules are that (i) we typically do not include auxiliary verbs, articles, demonstratives and temporal modifiers and (ii) we do not annotate parts of words.

**Baseline model configuration and results**

The early version of the model incorporated a relation classifier that also used the RoBERTa-Base transformer block as its primary component, but was structured as a single-label sentence-pair classification task. This relation classifier takes as input three components: the target sentence and string representations of the two spans whose relation is being predicted within the sentence. Each of these components is separated in the input using the reserved [SEP] token that is common to most NLP-related transformer models to mark sentence boundaries. The model then predicts the relation type between the target entities, or assigns no relation if the two entities are not linked by any of the relations defined in the annotation guidelines. To generate the training data for this model, we first generated all possible relations between the span tags in a sentence and then mark the pairs that exist in the gold standard with their respective label. All other pairs are treated as examples of non-relations.

Additionally, this baseline model predicted over a larger set of the annotation tags, many of which had low support and/or low inter-annotator agreement, and was trained on only 101 of the final 200 documents. As a result, the overall performance of the baseline model was not optimal and specific categories such as Perpetrator, Temporal_Frame performed poorly (Table S1). It was decided that the improvement in model performance achieved by not predicting low-performing entities outweighs the slight reduction in information from these annotations. Although the relation classifier performed well, this was calculated assuming perfect annnotation of the underlying entity tags that were the basis for the relation. In real circumstances, these types of entities (perpetrators and temporal_frames) proved as an aggregate to be less reliably identified, and subsequently, errors in identification propogate to the relation classifier, compounding overall error.

In comparing the performance of the baseline models to the clinical experts involved in the annotation task, it is challenging to establish a direct comparison for the relation extraction model. This is because when providing the inputs to the relation extraction model, we provide the span tags from the corpus, where span-level disagreements have been resolved during adjudication. Therefore, errors introduced by span-level disagreement (e.g. one annotator marking <shoved> and <father> with a Perpetrated_By relation between them, and another annotator marking <shoved> and <his father> with a Perpetrated_By relation between them) are not present in the model evaluation but are present in the human annotator error rate evaluation. As a result, the reported performance of the relation model is expectedly higher than the reported human performance, despite human performance generally being considered an upper bound of potential accuracy on the task. Computing the relaxed F1 metric ameliorates the issue of span-level disagreement but also significantly reduces the number of instances used to compute the human baseline, as any relation tags with span-level disagreements are excluded. The results of this earlier model are provided in Table S1, below.

**Table S1.** Per-label results of span-level (A) and relation classifier (B) for the baseline model.

*A. Span-Level model*

|  | precision | recall | f1-score | support | Human |
| --- | --- | --- | --- | --- | --- |
| **Event** | 0.549 | 0.719 | 0.622 | 320 | 0.660 |
| **Perpetrator** | 0.192 | 0.500 | 0.278 | 50 | 0.621 |
| **Substance** | 0.476 | 0.555 | 0.513 | 346 | 0.715 |
| **Symptom** | 0.629 | 0.754 | 0.686 | 2,665 | 0.706 |
| **Temporal_Frame** | 0.264 | 0.494 | 0.344 | 89 | 0.460 |
| **Macro Avg** | **0.422** | **0.604** | **0.489** | **3470** |  |

*B. Relation Classifier*

|  | Precision | Recall | F1 | Support |
| --- | --- | --- | --- | --- |
| **Perpetrated_By** | 0.732 | 0.703 | 0.717 | 101 |
| **Grounded_To** | 0.885 | 0.568 | 0.692 | 95 |
| **Macro Avg** | **0.806** | **0.638** | **0.705** | **196** |
