## Supplementary material for "Machine Learning in Psychiatric Health Records: A Gold Standard Approach to Trauma Annotation": TraumaAnnotationGuidelines-medrxiv

### TraumaML Annotation Guidelines

Version 1.0, January 2023.

### 1. Introduction

A vast body of evidence clearly indicates that a history of childhood (18 year old and under) trauma contributes to psychosis risk, substance use, self-harm behavior, and poor treatment outcomes. Yet systematic large-scale computational approaches to stratify subgroups of patients with and without trauma history and to examine the impact of childhood trauma on psychopathology or treatment outcome has yet to be developed. Annotation of patient records is a key step towards large scale, systematic queries of clinical text and training ML classifiers. The goal of the annotation is to mark up relevant clinical information present in the patient records (in our case: events, symptoms, temporal expressions, etc.), which would allow us to train NLP models to automatically extract this information from clinical text.

To our knowledge, annotation guidelines representing patients with psychosis and trauma-related features had yet to be developed. TraumaML fills at least part of this gap since it is designed for annotation of EHRs for patients with diagnoses of PTSD and psychotic disorders (schizophrenia, bipolar, schizotypal, delusional, or other non-mood psychotic disorder) at some point in patient history.

TraumaML includes extent tags that mark up a span of text and relation tags that indicate relations between extent tags:

| | |
|---|---|
| Extent Tags | EVENT, PERPETRATOR, SYMPTOM, SUBSTANCE, TEMPORAL_FRAME |
| Relation Tags | PERPETRATED_BY, GROUNDED_TO, SUB-EVENT |

### 2. Extent Tags

There are five extent tags that mark spans of text in the EHR.

| | |
|---|---|
| EVENT | The traumatic event that a patient has experienced |
| PERPETRATOR | The perpetrator of the traumatic event. |
| SYMPTOM | Symptom exhibited by the patient. |
| SUBSTANCE | Any text span that gives information about a substance use disorder. |
| TEMPORAL_FRAME | A temporal grounding for the traumatic event. |

#### 2.1. The EVENT Tag

The traumatic event that a patient has experienced.

Properties:

| | |
|---|---|
| Event_Type | The subtype of the traumatic event. One of “Sexual”, “Physical”, |

| | |
|---|---|
| | “Emotional” and “Other”. |
| Childhood_Trauma | Whether the event was a childhood trauma. Values: “True” and “False”. The default is “False”. |
| Factuality | Factuality or veridicity of the event. One of “Factual” (default), “Maybe” and “Unlikely”. |

The first three values for Event_Type are specific and should be used for text expressions referring to anything implicating that category name. The fourth category is used if there is no clear type for the event. Here are some instances of texts from records relating to the above categories (bold-faced and in red type, the category of the event is added between curly brackets):

- “The patient is a **-year-old gentleman who, four years ago, endured a [sexual assault] {Sexual} at the hands of his then girlfriend.”
- “… [raped] by several people at age ** {Sexual} {Childhood_Trauma}”
- “Reports significant h/o [physical] {Physical} and [verbal abuse] {Emotional} during childhood by stepfather, reports having frequent flashbacks, nightmares, dissociation…”

In the third case there is a conjunction and both parts are annotated separately. Annotators will annotate both [physical abuse] and [verbal abuse] as an event, which allows us to be precise on the categories. If we had annotated [physical and verbal abuse] as one event, then that would not have been possible. Note that in Brat “physical abuse” would be annotated as one event that has two fragments.

If a traumatic event is repeated in the text, then all instances are to be annotated.

Trauma has a broad meaning and includes medical conditions and diagnoses, these two are not considered traumatic events. For example, we do not annotate “traumatic brain injury”, “trauma”, “rupture of alveolar bleb” and “TBI” (Traumatic Brain Injury) in the following:

- “patient experienced traumatic brain injury following an accident”
- “trauma while scuba diving”
- “spontaneous rupture of alveolar bleb given his habitus”
- “TBI in <date> 2018”

Note that in the first two the injury was caused by some event (“accident” and “scuba diving”) that one might consider traumatic, but these are still just accidents and we do not annotate them as traumatic events.

- “patient experienced traumatic brain injury following an accident”
- “spontaneous rupture of alveolar bleb given his habitus”

However, in “pneumomediastinum caused by [tazer trauma]” we do annotate “tazer trauma” as an event because it was not an accident.

##### 2.1.1. Sub events

Traumatic events can have sub-events:

> “The patient is a **-year-old gentleman who, four years ago, endured a [sexual assault] at the hands of his then girlfriend. [….] He went to visit her one night. She [kneed him in the groin], [injected something into his penis], and then [sexually assaulted] him.

The second and third events can be considered sub events of the first event. We do not mark on those event that they are a sub event of the first event, but we do introduce a relation tag SUB-EVENT to deal with this (see section 3.3).

##### 2.1.2. Experiencer is not the patient

There are cases where a traumatic event is not experienced by the patient but by somebody else.

> “Per report, the patient's younger sister was also abused.”

We do not annotate traumatic events that were not experienced by the patient. However, witnessing a traumatic event (violence, hitting, physical assaults, etcetera) is an important clinical risk factor and falls under Adverse Childhood Experiences (ACEs) so we include them in the annotation as traumatic events of type “Other”.

> “The patient [saw her younger sister being abused]_{Other}.”

In the case above we annotate the entire span, not just the core event. Note that the event type is marked as “Other”, this is because it is not at all clear whether the abuse is sexual, physical or emotional.

##### 2.1.3. Factuality

Traumatic experiences in clinical notes may not be factual due to impairments in memory, delusions and/or exaggerations. In some cases, the text explicitly mentions that it is not clear whether the events did happen. We use a property on the EVENT tag named “Factuality”, which indicates the factuality or veridicity of the event. The default value is “Factual”, but this value can be overridden by the annotator with “Maybe” or “Unlikely”. The annotators should only use “Maybe” when the text specifically mentions words like “maybe/suspect/possible” and use “Unlikely” property if the clinical note explicitly brings up that the event is unlikely to have happened.

##### 2.1.4. Timing of the event

We use a binary feature *Childhood_Trauma*, which can be set to True of False depending on when the traumatic event occurred, with the default set to False.

##### 2.1.5. What span to annotate

Sometimes there are several choices of what span of text to include in the event, for example, with "her arm began shaking" we could annotate the entire span, which would be more informative, or we could just annotate "shaking" or maybe "arm began shaking", which would be more generalizable.

The general rule of thumb is to annotate the extent that is most informative and clinically relevant. For example, the "shaking" is the most important part in "her arm began shaking" and adding that it was the arm that was shaking does not add a lot of information or clinical relevance. On the other hand, "sexual" in "sexual assault" and "emotional" in "emotional trauma" bring in a lot of new relevant information because sexual and emotional assault are two of the event types that we recognize and the difference is clinically relevant. Note that clinically relevant differences are not always expressed as explicit event types. For example, with "military/combat trauma" we annotate the whole string, but we do not have a special subtype for this even though there is a large body of research on that kind of trauma. Note that if our annotation had been over a dataset from the VA then we might have added "military" as a subtype.

Two other general rules are that (i) we typically do not include auxiliary verbs, articles, demonstratives and temporal modifiers and (ii) we do not annotate parts of words.

These general rules also apply to the other extent tags.

Here are a couple of examples to make this more concrete:

- *"been [sodomized] by various objects including matches"*
  In this case the trauma is mostly in the sodomizing action, not in the specific objects used so we do not include anything but "sodomized".
- *"[physical aggression]" versus "physical [aggression]"*
  Use the longer extent since it really picks out a particular kind of aggression that is clinically relevant and helps to classify the kind of trauma we are looking at here.
- *"brother had a history of [physically hitting] mom and her"*
  Do not annotate parts of words. In this example, annotate "physically hitting" and not "physically hit".
- *"had [touched her without consent]"*

- Just annotate the main verb. In general, we do not annotate auxiliary verbs. This case is also interesting because we include the consent part, this is because touching by itself would not generally be considered traumatic.
- *"[childhood abuse]" versus "childhood [abuse]"*
  We just annotate "abuse", but "childhood" could be annotated as a temporal frame though. The same holds for "childhood [trauma]", "childhood [physical abuse]" and "childhood [sexual abuse]".
- *"intimate partner [violence]"*
  Use the smaller span since the left context can be annotated as a perpetrator.

#### 2.2. The PERPETRATOR Tag

The perpetrator of the traumatic event.

Properties:

| | |
|---|---|
| Perpetrator_Type | Indicates the relation between the perpetrator and the patient. Values: "Family-Member", "Colleague", "Partner", "Other-Known" and "Other-Unknown". |

Do not annotate people that were significant to the patient, but who were not perpetrators.

Examples:

- "Reports significant h/o [physical] and [verbal abuse] during childhood by [stepfather], reports having frequent flashbacks, nightmares, dissociation…"
- "The patient is a **-year-old gentleman who, four years ago, endured a [sexual assault] at the hands of his then [girlfriend]."
- "… stress at work due to an abusive [boss] and …"

#### 2.3. The SYMPTOM Tag

Any symptom that the patient exhibits or reports.

Childhood trauma is associated with a wide range of psychopathology and therefore we annotate not only PTSD trauma related symptoms, but also symptoms related to a broad range of psychopathology (psychosis, mood and anxiety). Note that symptoms may overlap across dx conditions (e.g., sleep problem may be symptoms of multiple psychopathology).

Properties:

| | |
|---|---|
| Not_Current_Symptom | Whether the symptom occurred recently. Values: “True” and “False”. |
| Negation | Indicates whether the occurrence of the symptom is negated. Values: “yes” and “no” (default). |

If a symptom is mentioned multiple times, then all instances should be annotated.

All symptoms should be annotated, be they past or current, so “cutting” in “history of cutting” would get tagged as a symptom. For past symptoms use Not_Current_Symptom. For example, with “Resolution of [suicidal thoughts] and [urges to harm self]” we annotated two symptoms and for each we add Not_Current_Symptom since the word resolution is saying it was there and now it's not.

Some examples of symptoms are:

- “avoidance,” “intrusive memories,” “flashbacks,” “nightmares,” “hypervigilance,” “PTSD symptoms,” “anxiety,” “tension,” “panic attacks,” “palpitations”, “interpersonal distress”
- “malaise,” “low appetite,” “weight gain/loss,” “sleeping all day,” “psychomotor retardation,” “psychomotor agitation,” “self-harm thoughts,” “suicidal ideation,” “attempted suicide,” “attempted to end his life”
- “delusions,” “hallucinations,” “responding to internal stimuli,” “loosening of associations,” “psychotic symptoms,” “mania,” “behavioral dysregulation”

Some examples of things that aren’t symptoms:

- General and common characteristics of people that are considered within the normal range are not annotated as symptoms, this includes “friendly and positive”, “bright and social”, “goal-directed”, “appetite is fair”, “insight and judgment were fair” and “poor coping skills”. None of those are psychiatric symptoms.
- “Decompensation” is not annotated because it is a way to state that a person is getting worse and it is not a symptom by itself.
- Annotate symptoms not dx. For example, Annotators will tag “anxiety” if it is mentioned as a symptom but will not tag it if it is mentioned as diagnosis. Other examples are “nervous tic disorder" and "history of tic disorder". Here symptoms are used in the context of a diagnosis so we do not annotate these spans, but “tic” or “tics” in isolation would be annotated.
- Don’t tag “Risk of harm to self and/or others” as it is a risk and not an actual symptom.

When annotating violent behavior, we often see sub symptoms like "took a scalpel and cut him", these very specific actions are not annotated. What we do annotate are more generic descriptions like "hurting people physically". Similarly, there are many variants of suicidal behavior. But that specific behavior is usually not used to describe a symptom. For example, in "patient attempted suicide by taking pills", it is "attempted suicide" that should be annotated as a symptom, not the entire phrase or "taking pills". Another example is "violent behavior due to [delusional thoughts]", where the thoughts are the symptom, not the violent behavior. On the other hand, if we have "patient overdosed on pills" then that specific action is all we have and we should annotate that.

When the text mentions "PRN" or "at pt's request" as in "PRN acute anxiety /agitation/ insomnia at pt's request" then do not tag "anxiety etcetera as symptoms because it doesn't mean that the patient has these symptoms. It is just saying medications will be offered should the patient have anxiety, agitation, or insomnia, in case they have it. It's not actually saying they do. However, when text says "patient was treated with <drug> for [anxiety /agitation/ insomnia]", then "anxiety", "agitation" or "insomnia" should be tagged.

Some other cases:

- In "scratch herself superficially as self-injury", tag "self-injury" as symptom. But "scratch herself superficially" will not be tagged because it is description of self-injury which has already been tagged. If a sentence contains only [scratch herself superficially] without mentioning self-injury, we will tag it. Similarly, we will not tag description of suicide attempts (e.g., jump from a bridge) unless the term -suicide attempt- is not there.
- Tag "frustration tolerance was low" as symptom of borderline personality disorder.
- Tag "disappointed in herself", as it captures concept of low self-esteem, worthlessness.
- Tag "characterological dysphoria" and "frustration tolerance was low" as symptoms to capture the concept of borderline personality disorder symptom.
- In "recall of the rape", tag "recall of the rape" as symptom to capture flashback. Also, tag "rape" as a traumatic event.

Symptoms can be annotated in cases like this:

> "[suicide attempts]/[self-harm] {Negation}: no past attempts"

The symptom occurs in a heading, which we usually do not annotate. But this is a very different header then something like "past history" which contains a generic name of a section, the current case is a very specific kind of header and will be annotated. Two more cases that look like this, but with more complex symptoms:

> "[speech]: [increased rates], [prosody], [amount] and [volume]"
> "[Affect]: [Increased range], [labile] ..."

In the first case we have four symptoms and each of them has "speech" as one of its fragments. The second case is a bit different since "labile" stands by itself as a symptom. "Increased range" will be bundled with "Affect" as a single symptom. Had the order been different ("labile" before

“Increased range”) then we would have had [Affect] and [Increased range] as two fragments of the one symptom.

##### 2.3.1. Implicit symptoms

Annotate symptoms that are stated in the notes, skip symptoms that are indirectly inferred from the sentences. For example:

> "She felt that she did not have any mental illness and did not need to be in the hospital."

These are not symptoms, but you could argue for "lack of insight" as a symptom. However, deducing this takes some mental effort, only annotate things that are in the text explicitly.

##### 2.3.2. Negation

Negation is important information for clinicians to rule out or rule in dx and treatment plans.

> She does not have flashbacks.
> She denies SI, HI, and access to a weapon.
> She denies any mania symptoms, AH/VH, paranoia, ideas of reference, thought insertion, thought withdrawal, or thought broadcasting.
> She denies anxiety.

There are two kinds of negation here, one where the text explicitly uses a negation marker (“not”) and one where the event is somehow qualified (“denies”). All events are assumed to be non-negated by default so negation should be marked explicitly:

> She does not have [flashbacks]_{Negation=True}.
> She denies [anxiety]_{Negation=True}.

This is different from cases where the negation is part of the symptom:

> … leading to him [not going to work] and [not paying his bills].
> (this is assuming here that the above are true symptoms)

Negation of symptoms are usually taken at face value but sometimes there are inconsistencies between what is observed and what is said. Consider the following example:

> … the patient denies [AH]_{Negation=True} …
> … however pt is noted to be [responding to internal stimuli].

We annotate what is said, so in this case we both annotate the patient’s statement and the remark by the clinician, even though they seem contradictory.

##### 2.3.3. What span to annotate

As with event extents, we ignore temporal and other qualifiers of the symptom and just annotate the core of the symptom, for example:

generalized [anxiety]
grandiose [delusions]
episodic [impulsivity]
general [malaise]

With "thoughts of wanting to harm herself" you annotate the whole string and not just "harm herself" since the latter did not happen and can thus not be a symptom. In addition, thoughts are an ideation and express more ambivalence than just "wanting to harm herself" and therefore the whole phrase has a different meaning than just the last four words.

*"history of complicated [withdrawal]"*
Do not include "complicated" since it just refers to the severity of withdrawal symptoms. In general, those kinds of qualifiers are not included.

"[visual hallucinations]" and "[auditory hallucinations]"
We annotate the entire span and not just the second word since the difference is clinically relevant sometimes sexual trauma is more likely to cause auditory hallucinations). In addition, these two are also often referred to as "AH" and "VH" where we would not just annotate the "H" (per the general rule of not annotating parts of words).

"[delusions] with [hyper-religious] themes"
This is annotated as one symptom with two fragments. Note that "hyper-religious delusions" is a flavor of "delusions" so the above does not include to separate symptoms.

"[feeling abandoned/hopeless/guilty/overwhelmed/suicidal]"
Include "feeling" since feeling abandoned, etcetera, is different from being abandoned. On the other hand, do not include "feeling" with "feeling suicidal" since when you feel suicidal you are in fact suicidal.

"[impulses to cut herself]"
Include "herself" in the symptom since the difference between self-injury and injuring other people is relevant.

"[worry that she might hurt herself]"
There is ambivalence here. The patient wants to hurt self, but also does not want to and worries she will. We annotate the whole string.

##### 2.4. The SUBSTANCE Tag

A substance is a drug, chemical, or other material (such as glue) that one is dependent on or uses habitually and that is often illegal or subject to government regulation. What we annotate is not the substance itself, but any text span that gives information about a substance use disorder (daily/problematic use, abuse, dependence, disorder and withdrawal, IVDU, detox, DT).

There are no properties on this extent tag.

Substances mentioned in lab results are not annotated. For example, we do not annotate "THC" or "benzodiazepines" in "Urine toxicology screens on arrival notable for THC and benzodiazepines."

Examples:

- relapsing with [IV heroin use]
- history of [complicated withdrawal]
- current and ongoing [daily heroin use]
- with 10+ [detox] admissions

We also do not annotate any frequencies associated with the disorder, as with the "detox" example above. On the other hand, we do annotate the full span in "[daily heroin use]" since the daily use is essential in determining the presence of a disorder.

"active [substance abuse]"
Do not include "active" since all it does is introduce temporal information.

##### 2.5. The TEMPORAL_FRAME Tag

Temporal information associated with an event, symptom or substance.

Properties:

| | |
|---|---|
| Temporal_Type | The kind of temporal frame. Values: "Age", "Period", "Time-of-life", "Event" and "Date". |

The temporal type reflects the various kinds of temporal information that can be captured:

| | |
|---|---|
| Age | The time of exposure as identified by the patient's age |
| Period | The period or duration of the exposure ("for two years", "after moving to Idaho", "after 9/11", "six months ago"). |
| Time-of-life | The time of exposure as identified by the patient's life phase, e.g., "childhood", |

| | |
|---|---|
| | or “adolescent”. |
| Event | Some event like “Korean war” or “9/11” |
| Date | The date of the exposure |

Examples:

- “Reports significant h/o [physical] and [verbal abuse] [during childhood] by [stepfather]"
- “The patient last used [alcohol] [six months ago].”

The “Temporal_Type” attribute on the tag distinguishes between the classes above, with values like age, duration, time-of-life, event, date.

Temporal frames can be quite complex, as in “since he was fourteen years old” and “the night after visiting the hospital”. In these cases, we annotate the entire span, not just the head of the phrase.

“[from ages 7-10]” versus “from [ages 7-10]”.
We will tag a shorter span “ages 7-10”.

“[as a child]” versus “as a [child]”
Annotate the shorter span.

### 3. Relation Tags

There are three relation tags that connect extent tags.

| | |
|---|---|
| PERPETRATED_BY | Connects an event to a perpetrator. |
| GROUNDED_TO | Connects an event to a temporal frame. |
| SUB-EVENT | Relation between an event and another event that it is part of. |

To reduce the strain of annotation and increase precision we typically only annotate relations between extent tags that occur close together, typically between tags within a sentence or at least between tags that are in the same paragraph.

#### 3.1. The PERPETRATED_BY Relation

| | |
|---|---|
| Arg1 | EVENT |
| Arg2 | PERPETRATOR |

Connects an event to a perpetrator.

> “Reports significant h/o [physical] and [verbal abuse] [during childhood] by [stepfather], reports having frequent [flashbacks], [nightmares], [dissociation]…”
> PERPETRATED_BY (physical … abuse, stepfather)
> PERPETRATED_BY (verbal abuse, stepfather)

In this case there are two traumatic events and each one of them is linked to the perpetrator.

##### 3.2. The GROUNDED_TO Relation

| | |
|---|---|
| Arg1 | The EVENT that is grounded to a time frame |
| Arg2 | TIME_FRAME |

Connects an event to a nearby temporal frame

> “Reports significant h/o [physical] and [verbal abuse] [during childhood] by [stepfather], reports having frequent [flashbacks], [nightmares], [dissociation]…”
> GROUNDED_TO (physical abuse, during childhood)
> GROUNDED_TO (verbal abuse, during childhood)

A conjunction like “physical and verbal abuse” is not annotated as one traumatic event but as two events (see the discussion on this in section 4). It does not show properly in the text above, but the two events are “[physical] and verbal [abuse]” and “[verbal abuse]”. Note that each of them is grounded to the same temporal frame, but it is possible for them to be linked to two different frames.

##### 3.3. The SUB-EVENT Relation

| | |
|---|---|
| Arg1 | EVENT, the sub event in the relation |
| Arg2 | EVENT, the event that includes the other event |

This relation is to deal with sub events. The example below has a couple of events that can be considered sub events of other events.

> “The patient is a **-year-old gentleman who, four years ago, endured a [sexual assault] at the hands of his then girlfriend. [….] He went to visit her one night. She [kneed him in the groin], [injected something into his penis], and then [sexually assaulted] him.
> SUB-EVENT (“kneed him in the groin”, “sexual assault”)
> SUB-EVENT (“injected something into his penis”, “sexual assault”)

All types of trauma can have sub events, not just sexual trauma. For example, with “physically abused by the father, being slapped or spanked or beat up” we a couple of sub-events of physical trauma.

### 4. Other Considerations

#### 4.1. Pre-annotations

Annotation is often done on data that is pre-annotated and therefore already includes some common symptoms. The symptom pre-annotation script may have annotated two conflicting spans. For example, it may annotate both “visual hallucinations” and “hallucinations.” Refer to the guidelines on what span to keep. In this case remove the annotation for “hallucinations” and keep the longer span.

It may also happen that a symptom like “persecutary delusions” is pre-annotated as two separate symptoms: “persecutory” and “delusions”. In these cases the two annotations should be combined into one annotation.

#### 4.2. Typos in the text

Typos are not uncommon. Annotate extents with typos if it is obvious what the typo is. If some text is so mangled that it is unclear that the extent is one of our defined tags, then do not annotate it.

#### 4.3. Structured data

We only annotate running text and ignore structured data. For example, nothing will be annotated in the following fragment (which is part of a larger section with structured data:

> Past Diagnoses: Bipolar D/O 1 w/ psychotic features; Schizoaffective D/O
> Past Hospitalizations: McLean (AB2) 11/2011 (manic and psychotic);
> Prior History of suicidality or suicide attempts: denied

However, any running text will be annotated:

> Hx of Significant Trauma: Yes
> Please describe:
> Per report, the patient was in an [emotionally] and [verbally abusive] relationship. She is no longer in that relationship. Also, per report, the patient was [sexually molested] at [the age of **] by a [** yo traveling artist ] while living in ***. Per report, the patient's sibling was also [abused].

On the other hand, non-generic headers with specific information should be annotated:

> [Suicide attempts]:

[Violence]:

Do not include the punctuation marker.

#### 4.4. Conjunctions

There are numerous examples of conjunctions that could be either annotated as on extent or as two extents where one of the extents would consist of two fragments. Examples are

“Suicide/Homicidal Ideation”
“Suicidal and homicidal Ideation”
“urges to harm herself or others”
“urges to cut and scratch”

Instead of annotating one extent we annotate two extents:

“[Suicide]/Homicidal [Ideation]” and “Suicide/[Homicidal Ideation]”
“[Suicidal] and homicidal [Ideation]” and “Suicidal and [homicidal Ideation]”
“[urges to harm herself] or others” and “[urges to harm] herself or [others]”
“[urges to cut] and scratch” and “[urges to] cut and [scratch]”

Other examples where we have one string which contains two annotations are:

“Violent/destructive thoughts”
“Suicide attempts/self-harm”
“opiate and alcohol use disorders”
“Suicidal thoughts: intent”
